# Changes in dengue virus transmission in Mexico: 2016-2023

**DOI:** 10.64898/2025.12.12.25342126

**Authors:** Oliver Simmons, Anna Vicco, Ruth A. Martínez-Vega, José Ramos-Castañeda, Ilaria Dorigatti

**Affiliations:** MRC Centre for Global Infectious Disease Analysis, School of Public Health, Imperial College London, London, United Kingdom; Facultad de Ciencias Médicas y la Salud, Universidad de Santander, Bucaramanga, Colombia; Centro de Investigaciones sobre Enfermedades Infecciosas, Instituto Nacional de Salud Pública, México

## Abstract

Mexico has experienced increased dengue incidence in recent years, and there are concerns that the changing climate may enhance dengue transmission in the future. Whilst previous studies have characterised spatial heterogeneities in the long-term average risk of dengue infection in Mexico, the extent to which dengue transmission changes year-to-year has not been investigated. Furthermore, the extent to which dengue’s four serotypes may differ in their transmissibility remains poorly characterised globally.

In this study we characterised the spatial and temporal variations in dengue transmission intensity, as defined by the force of infection, across 27 states in Mexico, by analysing the annual dengue cases reported to surveillance between 2016 and 2023 using established catalytic models and a new, serotype-specific extension of these models that characterises differences in the transmission intensity of the different serotypes.

We found evidence of large spatial, temporal, and serotype-specific heterogeneities in transmission intensity across Mexico. Serotype-specific force of infection estimates suggest that DENV-1 and DENV-2 have historically circulated at high levels across Mexico, with DENV-4 transmission intensity low throughout the period studied. Model estimated DENV-3 transmission intensity increased in some states in recent years, coinciding with large outbreaks.

This work quantifies serotype-specific differences in dengue transmission intensity using routinely collected case-notification data and demonstrates how extensive RT-PCR-testing and new rapid diagnostic tests capable of discerning infecting serotypes can help to better understand the contributions of different serotypes to transmission.

## Introduction

The annual number of suspected dengue cases in the Americas, whilst variable, has increased steadily since 2000^(1)^, with the estimated burden of dengue in the region 16.7 (95% confidence interval [CI] 15.6-17.7) million infections and 7.6 (95% CI 4.8-9.9) million febrile cases annually^(2)^. Dengue’s principal vector, the mosquito *Aedes aegypti*, thrives in urban areas, breeding in small containers of water, and an increasingly urban global population therefore puts a growing number of people at risk of the disease. Increases in temperature and changing rainfall patterns, direct results of climate change, already have and will continue to impact the global distribution of infection and disease in the future, with currently inhospitable arid and temperate regions possibly becoming suitable to *Aedes* mosquitoes, and therefore dengue^(3)^.

Changes in both urbanisation and climate are particularly acute in Mexico, where over 100 million people live in urban areas^(4, 5)^. Mexico’s dengue burden has historically been greater in coastal and tropical areas in the south of the country, where the disease is endemic or hyper-endemic, and lower in the northern-central part of the country corresponding roughly to the Mexican Altiplano, as transmission here is limited by the high altitude, especially in areas higher than 1700m above sea level (though exceptions to these generalities exist)^(6, 7)^. Despite these limiting factors, Mexico reported approximately 278000 cases of dengue in 2023, which represented the highest annual incidence in the country in the last decade (as recorded by the Pan American Health Organization (PAHO)), and the second highest number of cases in the Americas in 2023 (after Brazil)^(1)^. In 2024, dengue incidence in Mexico approximately doubled, with more than 558000 cases reported to PAHO. Previous studies have suggested that the burden of dengue in Mexico may worsen further over the coming years due to climate change^(7)^, although the extent and geographical distribution of the predicted change in dengue transmission differs between models^(3, 8)^.

Dengue virus (DENV) transmission in Mexico is currently highly cyclical, both on an annual basis, with cases peaking between August and November each year during the rainy season, and on a year-to-year basis, with large outbreaks occurring every 3 to 4 years^(1)^. This inter-annual pattern of dengue is most likely a result of the complex population immune profiles that result from infection with DENV^(9, 10)^, which is made up of four, often co-circulating, serotypes denoted DENV-1, DENV-2, DENV-3 and DENV-4. Upon infection, each serotype confers lifelong immunity to that serotype, and short-term immunity to heterologous serotypes, though estimates of the duration of this cross-immunity vary^(11)^. This complex relationship between immunity and disease also makes evaluations of dengue’s disease burden difficult. The extent to which primary and post-primary (including tertiary and quaternary) infections are reported to surveillance can also vary, not only due to differences in disease but also in surveillance^(12)^. The general underreporting of cases in Mexico (as in any other country) and local differences in the sensitivity of dengue surveillance (driven, for instance, by differences in the historical trends of local dengue transmission) imply that the true burden of dengue infection is not necessarily reflected by the magnitude of the reported data^(6, 13)^. The burden of dengue can, however, be estimated from the age-distribution of the reported case data through mathematical models.

The rate at which susceptible people become infected with DENV, known as the force of infection (FOI), can be estimated using a class of mathematical models referred to as catalytic models, first used to model DENV transmission dynamics by Ferguson et al.^(14)^, using either seroprevalence or case-notification data^(15–17)^. In previous work, Imai et al.^(17)^ used two seroprevalence studies published in Mexico to estimate the nationwide dengue FOI to be within the range 0.035-0.037, assuming life-long immunity (and within the range 0.090-0.098 assuming immunity decay). Subnationally, Rodriguez-Barraquer et al.^(18)^ used case notification data from 2000-2009 to estimate the FOI of dengue in 27 of Mexico’s 32 states, which ranged from 0.01 to 0.092 on average.

In light of the recent increases in dengue incidence in Mexico, in this study we aim to better characterise the past and recent transmission intensity of DENV across the country. To this aim, we present a descriptive analysis of publicly available line-list dengue data recorded in Mexico between 2016 and 2023 and characterise the average DENV transmission intensity over all serotypes in the country using established catalytic models developed by Imai et al.^(15)^ and O’Driscoll et al.^(16)^. Finally, building on the work of Imai et al.^(15)^ and O’Driscoll et al.^(16)^, we developed a new serotype-specific catalytic model of DENV transmission intensity to determine the transmissibility of each serotype, enabling us to investigate heterogeneities in the transmission and immunological drivers behind the dengue incidence trends observed in Mexico in the last 8 years.

## Methods

### Data

#### Epidemiological Data

In Mexico, the reporting of suspected dengue cases by healthcare centres is mandatory, and dengue cases are recorded by the healthcare system through the National Epidemiological Surveillance System (SINAVE). Access to anonymised line-list data on dengue cases in Mexico is available for 2020-2023 from the Mexican Ministry of Health (Secretaría de Salud) website^(19)^. The data for 2016-2019 are available upon request from the Instituto Nacional de Acceso a la Información (INAI), folio 330026924000362. The combined dataset comprised 833761 confirmed and probable cases, with information on the federal entity (administrative level 1, from here on referred to as “state”) and municipality (administrative level 2) of residence. We excluded international cases (those with state of residence listed as the USA, “other Latin-American countries” or “other countries”) from the analysis.

#### Population Data

Population data at administrative levels 1 and 2 for Mexico were reconstructed from the 2015 intercensal survey^(20)^ and from the 2020 census^(5)^. For each state, we calculated the change in population of each 5-year age group (*A*_1_, *A*_2_, …, *A*_16_, where *A*_1_ = 0-4 years old, *A*_2_ = 5-9 years old, …, *A*_15_ = 70-74 years old and *A*_16_ = 75+ years old) between 2015 and 2020 and reconstructed the yearly rates of population change, assuming that those whose age was listed as NA in the 2015 intercensal survey or in the 2020 census (0.07% of people in the 2015 intercensal survey and 0.22% (273,386/126,014,024) in the 2020 census) were distributed according to the age distribution of the rest of the population with known age in that year in each state. The population growth rate of age group *A_j_*, *r_j_*, *j*, ∈ {1, 2 …, 16}, was then used to estimate the population size of each age group in each state between 2015 and 2020, and we extrapolated the population size for 2021, 2022 and 2023 assuming the same growth-trend, using the formula:

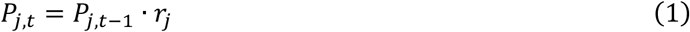

where *P_j,t_* is the population of age group *A_j_* in year t, t ∈ T = 2016, 2017, …, 2023}.

#### Serotype-specific case incidence

We reconstructed the number of serotype-specific cases from the total number of cases reported by state and the respective proportions of serotype-specific cases among hospitalised and non-hospitalised cases. To account for the possibility that hospitalised people may be more likely to receive an RT-PCR (reverse transcription polymerase chain reaction) test than non-hospitalised people, we generated serotype-specific case data by summing the reconstructed number of serotype-specific hospitalised cases (obtained by multiplying the proportion of hospitalised cases testing positive for a given DENV serotype amongst all the RT-PCR-positive hospitalised patients by the total number of hospitalised cases) and the reconstructed number of serotype-specific non-hospitalized cases (obtained by multiplying the proportion of non-hospitalised cases testing positive for a given DENV serotype amongst all the RT-PCR-positive non-hospitalised cases by the total number of non-hospitalised cases).

We assumed that the serotype distribution of the cases with unknown hospitalisation status was the same as was observed among non-hospitalised cases. Building on the available age-distribution of serotype-specific case data (Figures S5 and S6, Supplementary Information), we assumed that the observed serotype proportions were constant over all ages. We only reconstructed the serotype-specific case incidence in a given state for years that had more than 10 RT-PCR-positive test results, and we assumed that a DENV serotype was present in a state if there were more than 10 RT-PCR-positive results for that serotype in a at least one of the modelled years (2016-2023 inclusive).

### Mathematical Models

#### Non-serotype-specific models

To estimate the FOI in each of Mexico’s 32 states over the 8 years of data (2016-2023 inclusive), we used both time-constant and time-varying catalytic models. The time-constant catalytic model (Model 1) was developed by Imai et al.^(15)^ and assumes that the per-serotype FOI (denoted λ in the model) is constant throughout the 8 modelled years and historically prior to 2016. The explored four variants (Model 2-5) of the time-varying catalytic model developed by O’Driscoll et al.^(16)^ allow the per-serotype FOI, denoted λ_’_, to vary each modelled year and assume a constant historical per-serotype FOI prior to 2016. Both the original time-constant and time-varying models assume that the four DENV serotypes are equally transmissible and that tertiary and quaternary cases are not symptomatic, and hence not reported.

For all models, the reported incidence of dengue cases in age group 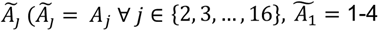 year olds (as we removed those under 1 year old from the analysis)) in year *t*, *D_j,t_*, can be described by equation (2):

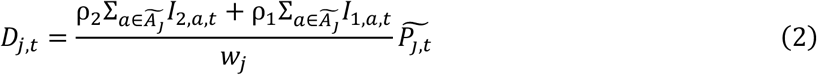

where *I*_1,*a,t*_ is the annual incidence of primary dengue infection in age *a* in year *t*, *I*_2,*a,t*_ is the annual incidence of secondary dengue in age *a* in year 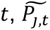 is the population of 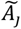 in year 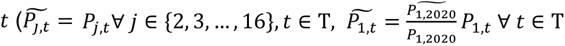, where *P*_1,2020_ and 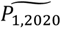 are obtained directly from the 2020 intercensal survey) reconstructed as defined in equation (1), ρ_2_ is the reporting rate of secondary cases, ρ_1_ is the reporting rate of primary cases, and *w_j_* is the width of age group 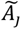 (e.g. for 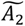 the age group containing 5-9 year olds, 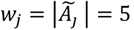).

Model 1 assumes that that the reporting rate of primary infections relative to secondary is smaller or equal to the reporting rate of secondary infections. We varied assumptions about the relative reporting rate of primary infections compared to secondary infections and the rate of reporting in children under 15 years compared to those over 15 between Models 2-5 (see Supplementary Information Table S3), and used the deviance information criterion (DIC)^(21)^ for model selection. In brief, Model 2 followed the same assumptions as in O’Driscoll et al.^(16)^; Model 3 allowed the reporting rate of primary infections to be greater than that of secondary infections (though it could also still be lower if that provided the better fit); Model 4 allowed different reporting rates for children under 15 years old compared to those older than 15 years, to reflect potential differences in the diagnosis and reporting of dengue cases through paediatricians; and Model 5 included the assumptions used in both Model 3 and Model 4, i.e. differential reporting rates for children under 15 years and no restriction on the relative reporting of primary vs secondary infections. We assigned *Normal(0,1)* prior distributions, constrained to be positive and below 1, to all parameters in Models 1-5, apart from the scaling factor used in Model 5, *χ*, which was assigned a *Normal(1,1)* prior distribution (constrained to be positive).

#### Serotype-specific model

We developed a variation of Model 5 to allow serotype-specific time-varying FOIs, denoted as 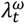 (ω ∈ ∈ Ω = {1, 2, 3, 4} denoting the 4 serotypes, *t* ∈ T denoting the year, with T varying depending on the data available). Equations (3-11) describe this new model (Model 6), where S(a, t) denotes the proportion of susceptible population of age *a* at the end of year *t*, *MO*(*a*, *t*, ω) the proportion of the population monotypically infected with dengue serotype ω at age *a* at the end of year *t*, *MU*(*a*, *t*) the proportion of the population multitypically infected at age *a* at the end of year *t*, *I*_1_(*a*, *t*, ω) the annual primary incidence of dengue serotype ω in age *a* in year *t* and *t*, *I*_2_(*a*, *t*, ω) the annual secondary incidence of dengue serotype ω in age *a* in year *t*. In equations (3-11), 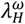 is a constant historical per-serotype force of infection that is used to reconstruct the initial immunity profile of the population at the start of the time-series data, and Ω denotes the set of DENV serotypes.

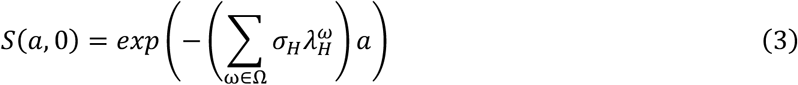

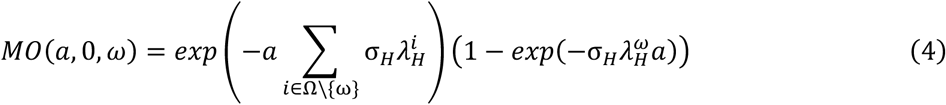

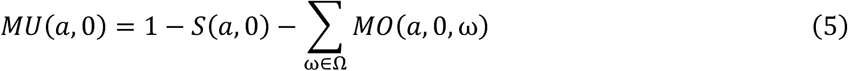

for *t* ∈ T, ω ∈ Ω, a ≠ 0

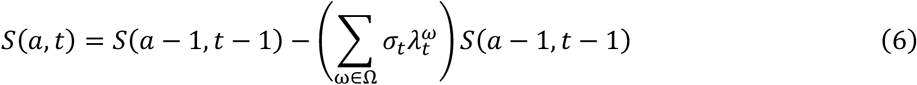

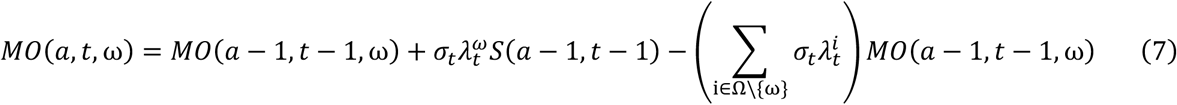

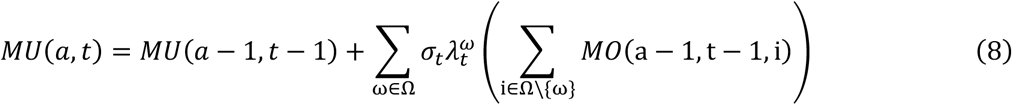

for *t* ∈ T, ω ∈ Ω, ∀*a*

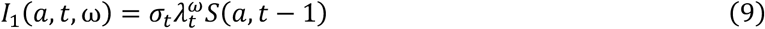

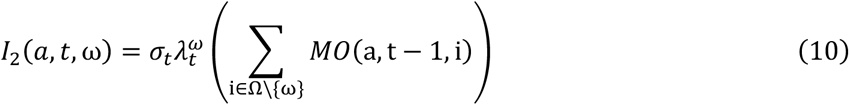

The serotype-specific number of cases reported in age group 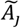 during year *t* for serotype ω, 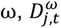, was modelled as:

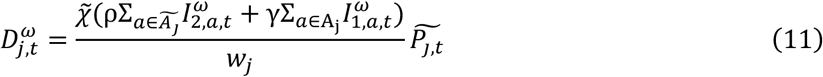

where ρ represents the reporting rate of secondary cases, γ the reporting rate of primary cases, 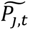 the population of age group 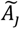 in year *t* (as defined above), *w_j_* the width of age group 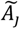 and where 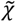 is a multiplier that allows children under 15 to report at a different rate than people older than 15 (i.e. 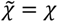, which we estimate, and 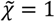 otherwise).

We assigned a Dirichlet prior distribution to 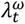 at each timestep and included a scaling factor σ_t_ (σ*_H_* for the historical 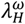) to allow the total FOI to be below 1, which was assigned a *Uniform(0,1)* prior distribution. Prior distributions for the other parameters were the same as for Model 5.

#### Model Fitting

We fitted our models to either age-stratified case-notification data (Models 1-5) or reconstructed age-stratified serotype-specific case incidence data (Model 6), excluding cases in children younger than 1 year old due to maternally transferred immunity^(22)^. We assume that the total number of cases (over the study period for Model 1 and over each year for Models 2-5) was Poisson distributed and that the age-specific number of cases was multinomially distributed. Similarly, for Model 6, we assume that the yearly total number of cases (over all age groups) for each serotype was Poisson distributed, and that for each serotype the yearly age-specific distribution of cases was multinomially distributed.

All models were fitted to the data using Hamiltonian Monte Carlo, a Markov Chain Monte Carlo method implemented in *rstan*^(23)^. For each state, models were run using 3 chains, each with 6000 iterations, of which the first 1000 were considered warm-up. The chains were considered to have successfully converged for all parameters if the diagnostic value *Rhat* was below 1.05 and the bulk effective sample size (ESS) was over 300, as per Stan guidance^(23)^. We excluded 5 states from our modelling analyses: Baja California, Ciudad de México, Chihuahua, Tlaxcala, and Zacatecas, as there was no evidence of sustained transmission over the study period in these states.

### Data Availability

Data cleaning, manipulation, analysis and visualisation were conducted in R version 4.4.1^(24)^ using the packages *tidyverse*, *sf*, *MetBrewer, patchwork* and *binom*. All models were coded and run using Stan^(23)^ through *rstan* version 2.35.0.9000. The code and data are publicly available at: https://github.com/mrc-ide/Overall-and-Serotype-Specific-Dengue-Transmission-Trends-in-Mexico-2016-2023

## Results

### Descriptive Analyses

Of the 833630 probable or confirmed dengue cases in Mexico between 2016-2023, 53.9% (449127) were in females, and the reported incidence among females was significantly higher than amongst males (874 vs 786 probable or confirmed case per million people per year, p-value < 0.001, chi-square test). Reported dengue incidence varied over time, with the highest incidence rates during the outbreak years of 2019 and 2023 (1.86 and 1.73 cases per 1000 people, respectively) (Figure 1a). Nationally, the median age of dengue cases ranged from 23 years in 2018 to 28 years in 2020, and the median age of reported cases was higher in females (27 years) than males (23 years). States in the south of the country showed a lower median age of reported cases than states in the centre and north (Figure 2a and Table S1, Supplementary Information).

**Figure 1:**
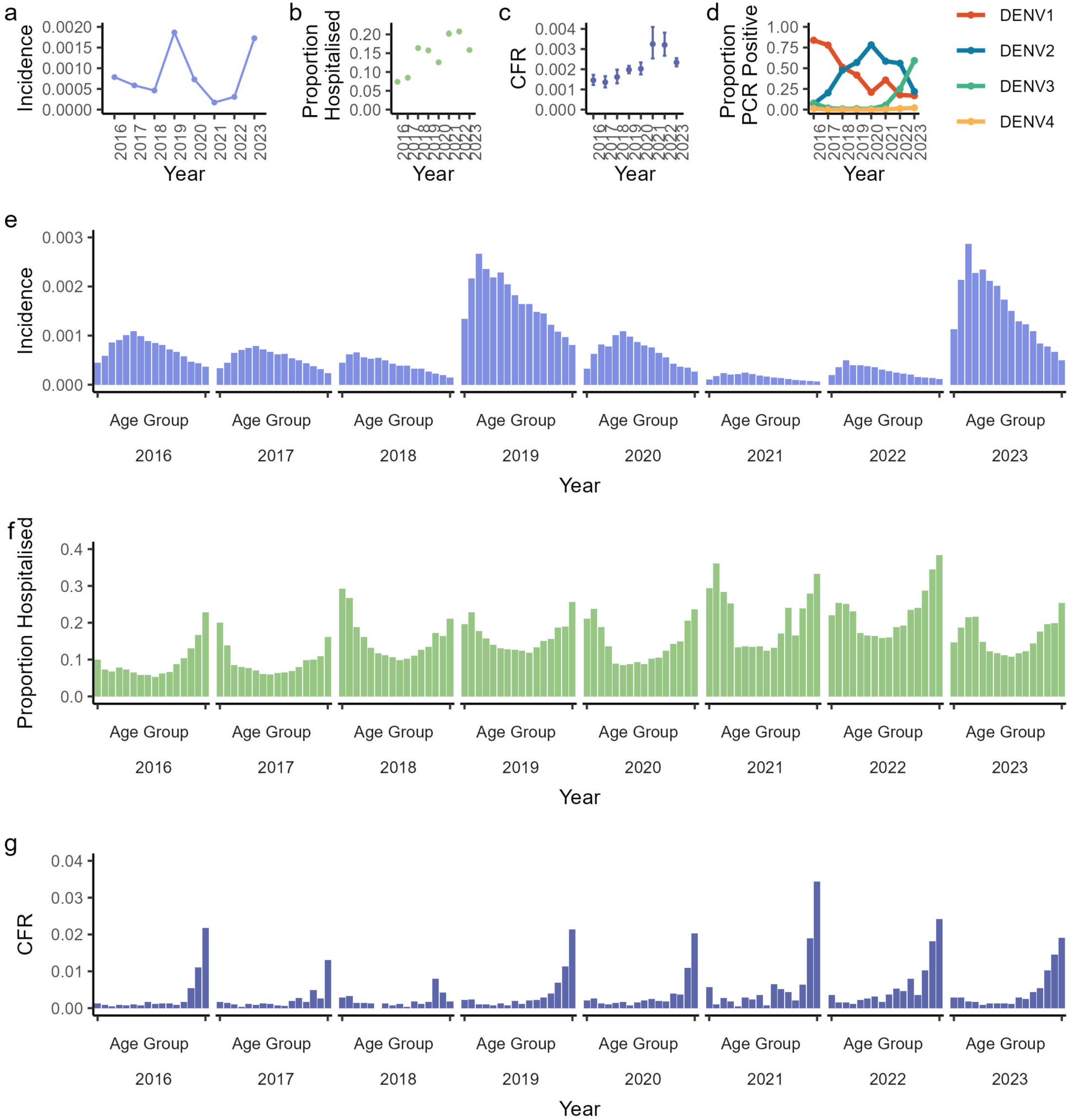
National incidence of reported probable or confirmed cases, hospitalisation, and case fatality ratio (CFR). (a) Yearly reported case incidence; (b) yearly proportion of cases hospitalised among those reported (c) yearly CFR, calculated as the yearly number of dengue deaths among those with dengue divided by the yearly number of reported dengue cases, (d) proportion of serotype-specific cases positive for each dengue serotype, (e) yearly age distribution of case incidence, (f) yearly and age-specific proportion of cases hospitalised and (g) age-specific CFR by year. Ages were split into 5-year bands (0-4 years, 5-9 years, up to 70-74 years and 75+ years). Plots b and c include the 95% exact binomial confidence interval for each point.

**Figure 2:**
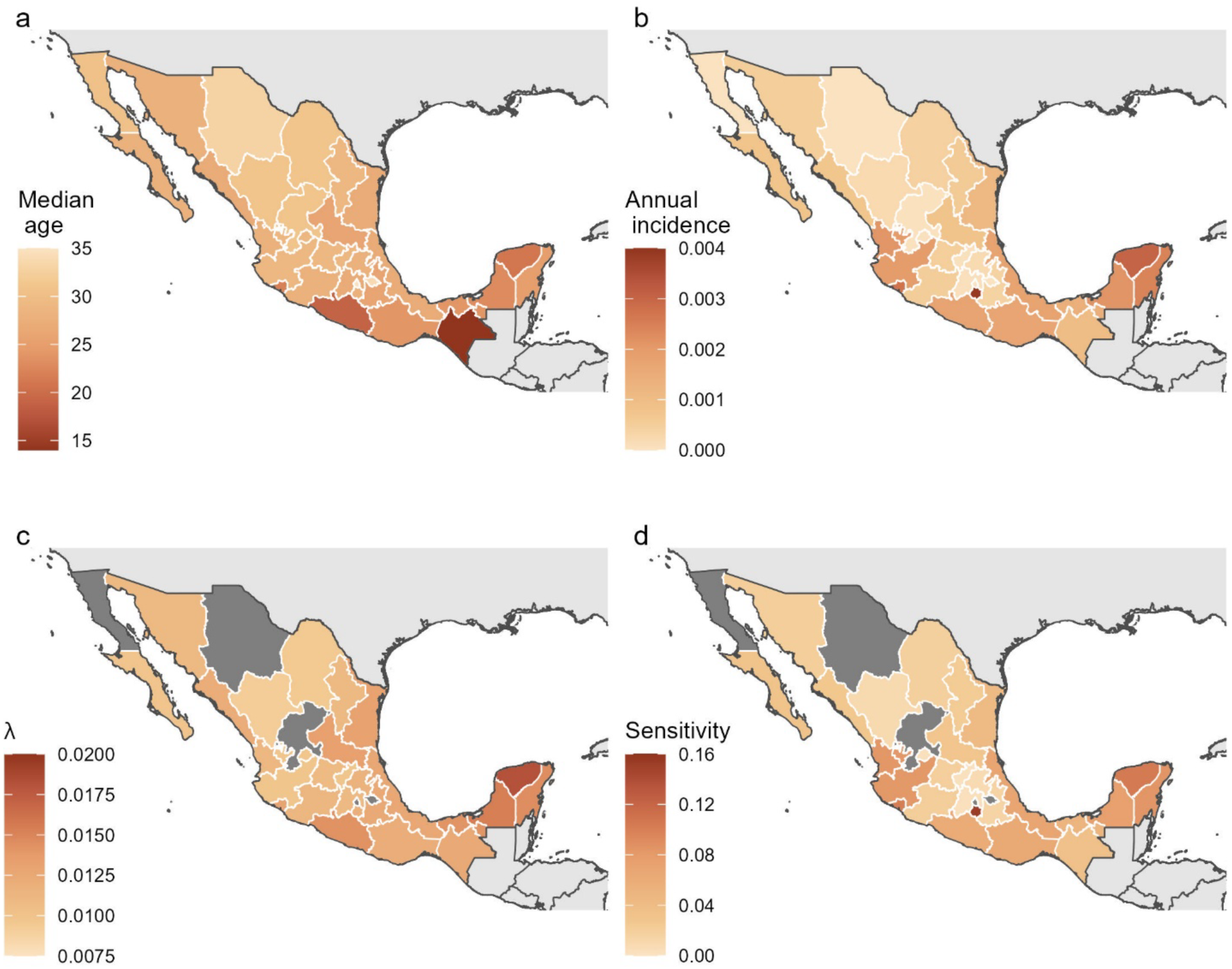
Spatial heterogeneities in the age of cases, incidence, force of infection and sensitivity of surveillance during 2016-2023. Median age of reported probable/confirmed dengue case by state (a), dengue incidence per person per year by state over the period 2016-2023 (b), median estimated time-constant per-serotype FOI (λ) for each of the 27 modelled states in Mexico between 2016-2023, estimated using a non-serotype specific model (Model 1) fitted to line-list case data of probable and confirmed dengue cases (c) and estimated sensitivity of surveillance, measured as the number of reported probable/dengue cases divided by the median expected number of infections from Model 1 (d).

The severity of dengue during the study period varied by age and location. At the national level, the proportion of hospitalised cases by age followed a “U” shape, with a greater proportion of hospitalised cases among the young and the elderly (Figure 1f). Nationally, over the entire study period, 14.2% (118738/833577; 95% exact binomial CI 14.2-14.3%) of confirmed and probable cases were hospitalised. Hospitalisation rates were highest in the oldest age groups and significantly higher among males 75 years old and over than among females in the same age group; 27.6% (1297/4698; 95% CI 26.3-28.9%) compared to 22.2% (1160/5216; 95% CI 21.1-23.4%; p-value < 0.001, chi-squared test). Geographically, states in the south and north-east of Mexico tended to admit the highest proportion of dengue cases to hospital (see Figure S2), with the greatest proportion of probable or confirmed cases hospitalised in Chiapas (30.6%; 13671/44605; 95% CI 30.2-31.1%), Tlaxcala (26.8%; 26/97; 95% CI 18.3-36.8%) and Guerrero (24.9%; 12311/49388; 95% CI 24.5%-25.3%).

The overall case fatality ratio (CFR) during the study period was 0.203% (95% CI 0.194%-0.213%), with 1696 deaths reported among those with confirmed or probable dengue. CFRs were highest in 2021 (0.325%; 71/21866; 95% CI 0.254-0.409%) and 2022 (0.321%; 127/39611; 95% CI 0.267-0.381%) (Figure 1c). The highest CFRs were observed in older age groups (Figure 1g), and, mirroring trends seen among hospitalisation rates, older males had a higher CFR than older females. Males over the age of 75 had the highest CFR of any group, 2.49% (117/4698; 95% CI 2.06-2.98%), while the lowest CFR was amongst males aged 20-25 (0.0733%; 30/40942; 95% CI 0.0495-0.105%). Regionally, the highest CFRs were recorded in the lower burden states in north-east Mexico such as Baja California (0.591%; 3/508; 95% CI 0.122-1.72%), Sonora (0.524%; 62/11843; 95% CI 0.402-0.671%) and Chihuahua (0.929%; 3/323; 95% CI 0.192-2.69%) (see Figure S2, Supplementary Information).

Dengue incidence was highly regionally heterogeneous during the reporting period (Figure 2a). 29.5% of the overall reported dengue case burden between 2016 and 2023 was concentrated in Jalisco and Veracruz, Mexico’s third and fourth largest states by population, which were worst affected during the 2019 outbreak, reporting, respectively, 74023 and 43611 dengue cases that year. The 2023 outbreak struck states further to the south, such as Yucatán, Morelos, Quintana Roo and Campeche, which reported 44646, 31946, 8934 and 19825 cases, respectively, corresponding to incidences of 18.1, 15.9, 9.45, and 9.39 cases per 1000 people (Figure S1). Morelos, Yucatán and Colima reported the highest dengue incidences between 2016 and 2023 (3.89, 3.03 and 2.72 cases per 1000 people per year, respectively) (Figure 2b), while the lowest dengue burdens were seen in Tlaxcala and the northern states of Chihuahua, Baja California, and Zacatecas, which reported 97, 323, 508 and 548 confirmed or probable cases of dengue over the entire 8-year period, respectively. These represented the lowest per-capita reported incidences in the country, along with Ciudad de México.

Among all cases, 85529 (10.3% of all probable or confirmed cases) tested RT-PCR positive for one of the four DENV serotypes between 2016 and 2023, with 35.6% (30473) of these positive for DENV-1, 45.5% (38894) for DENV-2, 18.2% (15535) for DENV-3 and 0.7% (627) for DENV-4. Post-2021, DENV-3 circulation steadily increased in the south of Mexico, particularly in states in the Yucatán peninsula such as Yucatán, Quintana Roo and Campeche, where DENV-3 accounted for 95.6% (4551/4759), 94.8% (2293/2420) and 98.5% (603/612) of all RT-PCR-confirmed DENV cases in 2023, respectively (Figure 3).

**Figure 3:**
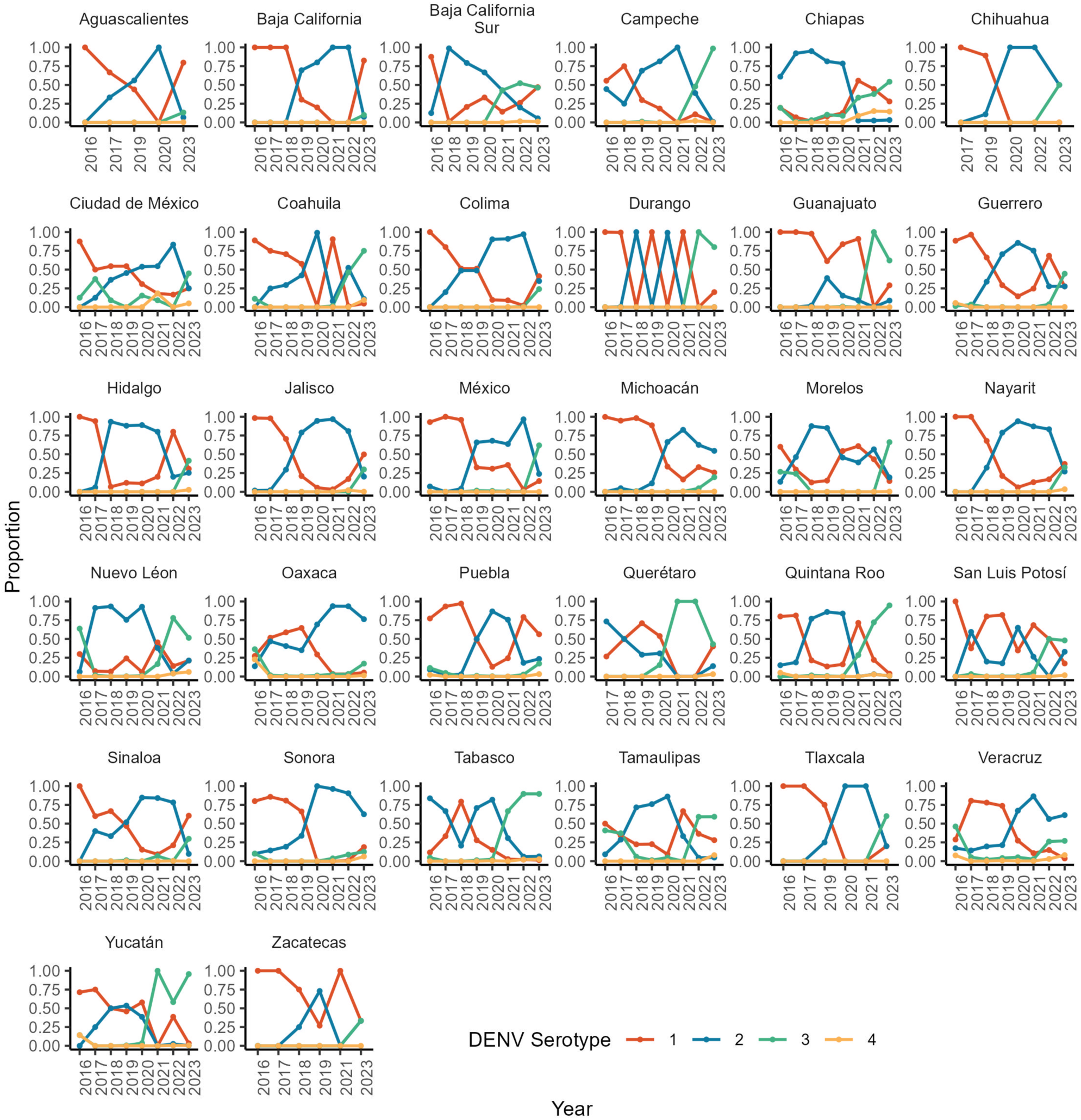
Spatiotemporal heterogeneity of DENV serotypes. Proportion of RT-PCR-positive dengue patients positive for each of the four serotypes of DENV. Each panel represents a di>erent state (by year), and each serotype is shown in a di>erent colour.

### Non-serotype-specific models

#### Time-constant model

Figure 2c shows the per-serotype FOI (λ) estimates from the time-constant non-serotype-specific model (Model 1) by state, showing large variations from 0.0092 (95% Credible Interval [CrI] 0.0086-0.0097) in Durango to 0.0182 (95% CrI 0.0179-0.0184) in Yucatán. Overall, our results suggest a higher FOI in the south of the country, particularly in the Yucatán peninsula. There was also substantial variation in the estimated secondary reporting rates (ρ) and the estimated reporting rates of primary cases relative to secondary reporting rates (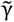), with ρ varying from 0.0051 (95% CrI 0.0046-0.0058) in Querétaro to 0.3245 (95% CrI 0.3207-0.3283) in Morelos, and 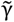 from 0.0003 (95% CrI: 0.00001, 0.0018) in Coahuila to 0.9920 (95% CrI 0.9604-0.9997) in Chiapas. Low estimated secondary reporting rates were often associated with low estimates of λ. We therefore estimate that the sensitivity of surveillance in Mexico between 2016-2023, defined as the total number of reported probable and confirmed cases divided by the total model-derived number of infections, varied from between 0.210% (95% CrI 0.196-0.228%) of infections detected in Aguascalientes to 15.3% (95% CrI 15.2%-15.4%) of infections detected in Morelos (Figure 2d).

#### Time-varying models

According to the DIC, the time-varying-FOI model variants that allowed differential reporting for children under 15 years old (Model 4 and Model 5) outperformed those that did not in 26 out of the 27 modelled states (see Table S2, Supplementary Information). Since Model 5 performed marginally better than Model 4 in 16 states, and considerably better in one (Chiapas), we only present the results from Model 5 below. Model 5 estimated lower reporting rates for primary infections compared to secondary infections in all 27 states, in line with the assumption used in Model 4, and both Model 4 and Model 5 estimated the reporting rate of children under 15 to be higher than that of people older than 15 in all states except 6 (Baja California Sur, Coahuila, Durango, Guanajuato, Michoacán and Querétaro).

Figure 4 shows the median per-serotype FOI estimate, λ_;_ (by definition assumed to be the same for all four serotypes) from Model 5, which is highly heterogeneous both in space and time. For instance, in Morelos, which is the state with the highest per-capita number of detected dengue cases over the reporting period, the median estimated λ_;_ varied from 0.0223 (95% CrI 0.0204-0.0234) in 2016 to 0.0057 (95% CrI 0.0052-0.0063) in 2022, before rising again to 0.0909 (95% CrI 0.0834-0.0985) in 2023. During 2023, the most recent year modelled, estimates of λ_;_ ranged from a median of 0.0003 (95% CrI 0.0002-0.0004) in Nuevo León in the north-east of the country and 0.00030 (95% CrI: 0.00029-0.00032) in Jalisco in the west, to 0.1152 (95% CrI 0.1085-0.1226) in Guerrero and to as high as 0.2495 (95% CrI 0.2475-0.2500) in Yucatán and 0.2488 (95% CrI 0.2436-0.2500) in Campeche in the south-east.

**Figure 4:**
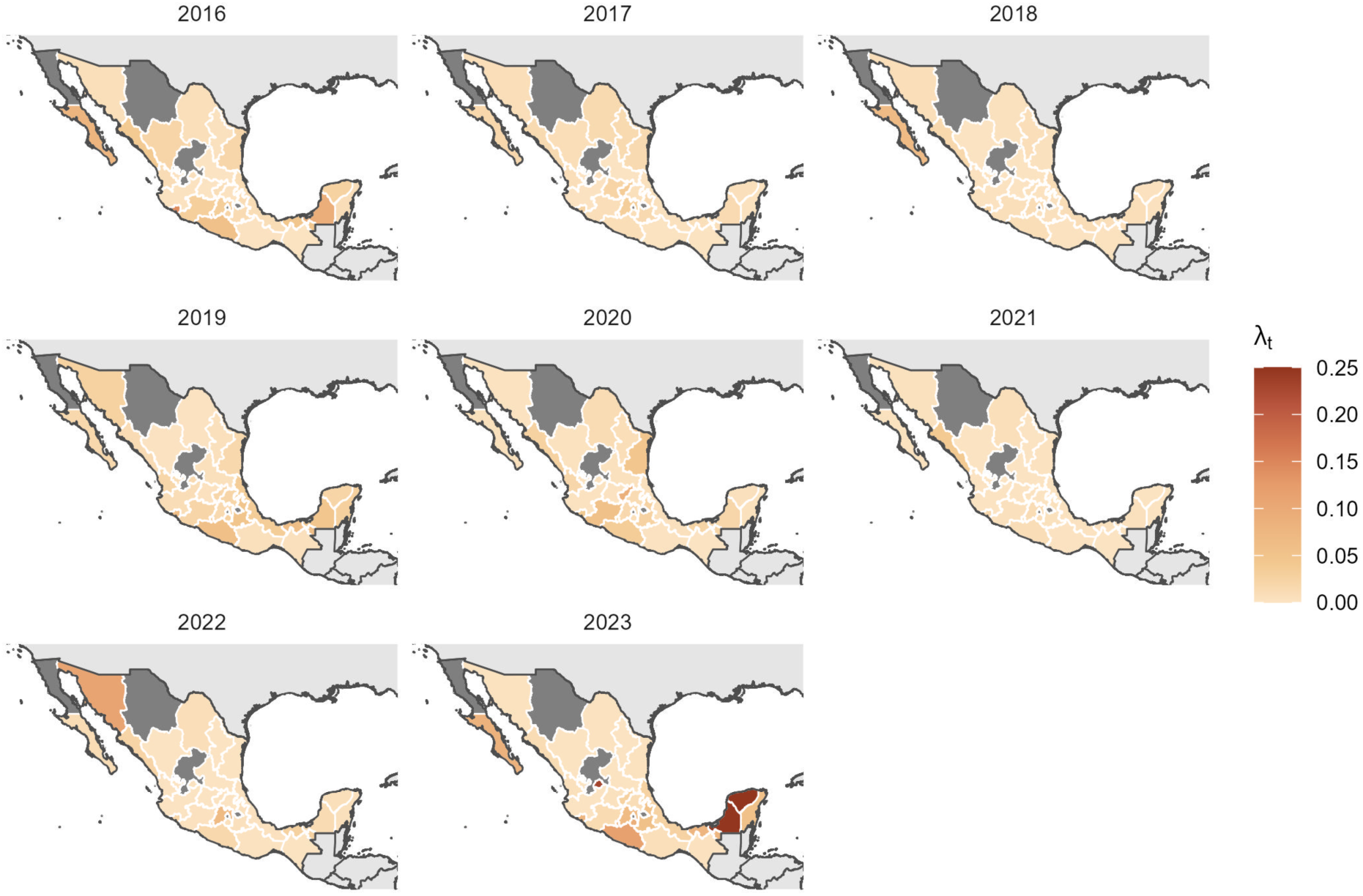
Spatiotemporal heterogeneity in annual state-level per-serotype FOI. Map of the median estimated time-varying per-serotype FOI for each of the 27 modelled states in Mexico between 2016-2023, obtained using a non-serotype specific model (Model 5), which allows for variable reporting for children under 15, fitted to line-list case data of probable and confirmed dengue cases.

Our model estimates that 10 out of 27 states had their highest dengue FOI during 2023. The states that saw the highest estimates of λ_;_ over the 8-year period were not, however, geographically clustered: Yucatán, Campeche and Guerrero in the south; Aguascalientes, Querétaro and Morelos in the centre; Sonora and Baja California Sur in the north; and Colima in the west. In contrast, lower FOI estimates are clustered in the centre of the country, especially in Guanajuato, San Luis Potosí, and Durango.

#### Serotype-specific Model

The limited number of RT-PCR-positive cases in the data meant that we only were able to model all 8 years for 11 of the 27 states, and a varying number of years for the other 17 states. We fitted the serotype-specific model in 9 states that had all four DENV serotypes cocirculating, 14 states in which DENV-1, DENV-2 and DENV-3 were co-circulating, and three states that only experienced circulation of DENV-1 and DENV-2. Durango was not included in the serotype-specific analysis, as only DENV-1 was circulating throughout the period studied. We were able to reconstruct the estimated age-distribution of the serotype-specific data (see Methods) using the serotype-specific model in 23 out of 26 states.

We found that the serotype with the greatest transmission intensity varied between states within a given year; in 2023, for example, DENV-1 had the largest FOI estimate in 6 states, DENV-2 the highest in 5 states and DENV-3 in 9 states. Each serotype’s transmission intensity also varied by year, with the highest estimated DENV-1 FOIs in 2016, 2019 and 2023, and the highest DENV-3 FOI estimates in either 2022 or 2023, clustered regionally in the south and to a lesser extent in the north of the country (Figure 5a). The years in which DENV-2 transmission intensity was greatest varied by state, and DENV-4 had the lowest estimated transmission intensity of the four serotypes, with its median estimated FOI only rising above 0.01 in two years in two states (Guerrero in 2016 and Chiapas in 2023). Low estimated DENV-4 FOIs were not associated with a high historical DENV-4 FOI estimate (Figure 5b).

**Figure 5:**
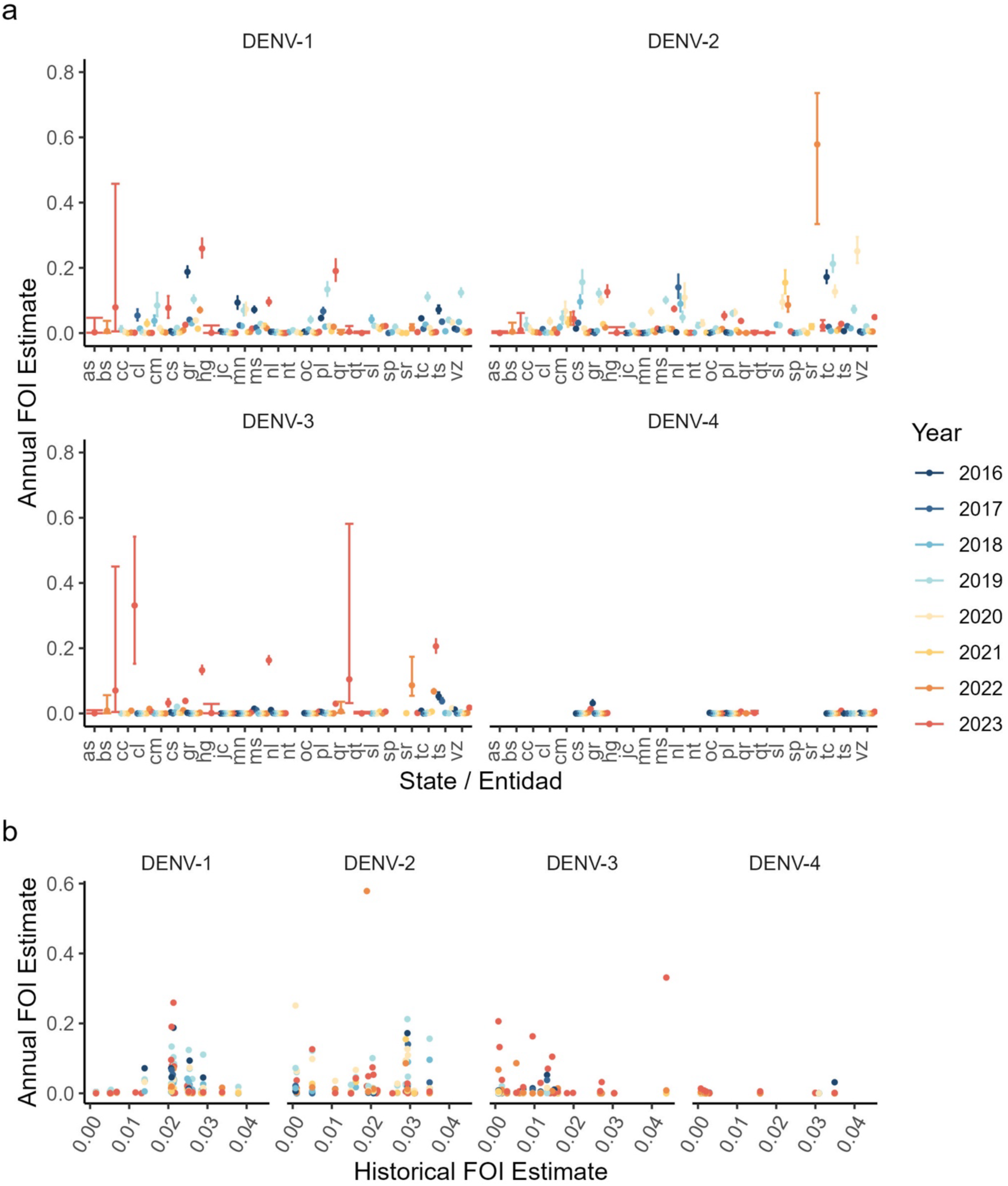
Annual serotype-specific force of infection (FOI) estimates by state. Serotype-specific FOI by state (a) and by modelled historical force of infection (b), using Model 6. State abbreviations: as – Aguascalientes, bs – Baja California Sur, cc - Campeche, cl – Coahuila, cm - Colima, cs – Chiapas, gr – Guerrero, hg – Hidalgo, jc – Jalisco, mn -, ms – Morelos, nl – Nuevo Léon, nt – Nayarit, oc – Oaxaca, pl – Puebla, qr – Quintana Roo, sl – Sinaloa, sp – San Luis Potosí, sr - Sonora, tc – Tabasco, ts – Tamaulipas, vz – Veracruz.

Figure 6 shows the modelled serotype-specific FOI estimates for Oaxaca, as well as the model’s fit to the estimated serotype-specific incidence, and the data used to estimate this incidence (the equivalent figures for each of the other 22 states are reported in Figures S7-S28).

**Figure 6:**
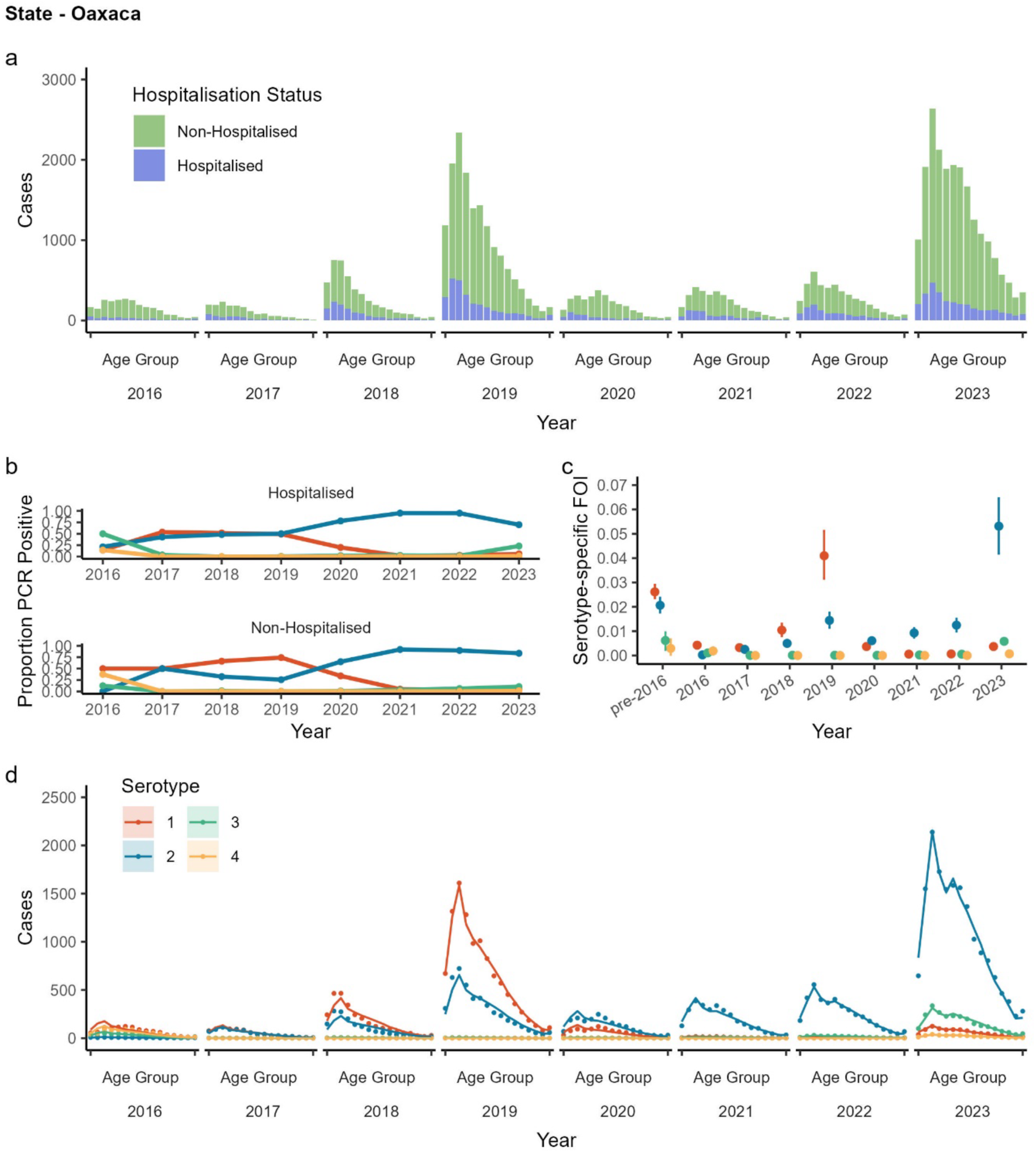
Data and outputs of the serotype-specific model (Model 6) for the state of Oaxaca. (a): Number of hospitalised (blue) and non-hospitalised (green) dengue cases per year per five-year age group 0-4, 5-9, …, 70-74, 75+.(b): Proportion of positive RT-PCR-test results positive for each serotype of dengue each year (DENV-1 red, DENV-2 blue, DENV-3 green and DENV-4 yellow), among hospitalised patients (top) and non-hospitalised patients (bottom). (c): Model estimated median serotype-specific FOI and 95% credible intervals for each serotype of DENV modelled (colouring the same as in plot b) for each year, 2016-2023. (d): Model fit (solid lines) to estimated serotype data imputed into the model (dots) by year and by serotype (colours same as before). Imputed data was generated by combining the proportions positive for each serotype among the hospitalised and non-hospitalised patients each year with the case data on annual probable/confirmed hospitalised and non-hospitalised patients.

The amount by which the overall yearly FOI estimates generated by the serotype-specific model (i.e. the sum of all the serotype-specific estimates for a given year) differ from those generated by the non-serotype-specific model (Model 5) varies for states fitted to more than five years of data (13/23 states). For the majority of these states, the difference between the total FOI estimated from the serotype-specific and the FOI from the non-serotype specific model is small (Figure 7). However, a large difference in the serotype-specific and non-serotype-specific overall FOI estimates was observed in Colima, Chiapas, Nuevo León, and Oaxaca, and states where fewer years of serotype-specific data were available (e.g. see Figure 7 and Figures S3 and S7).

**Figure 7:**
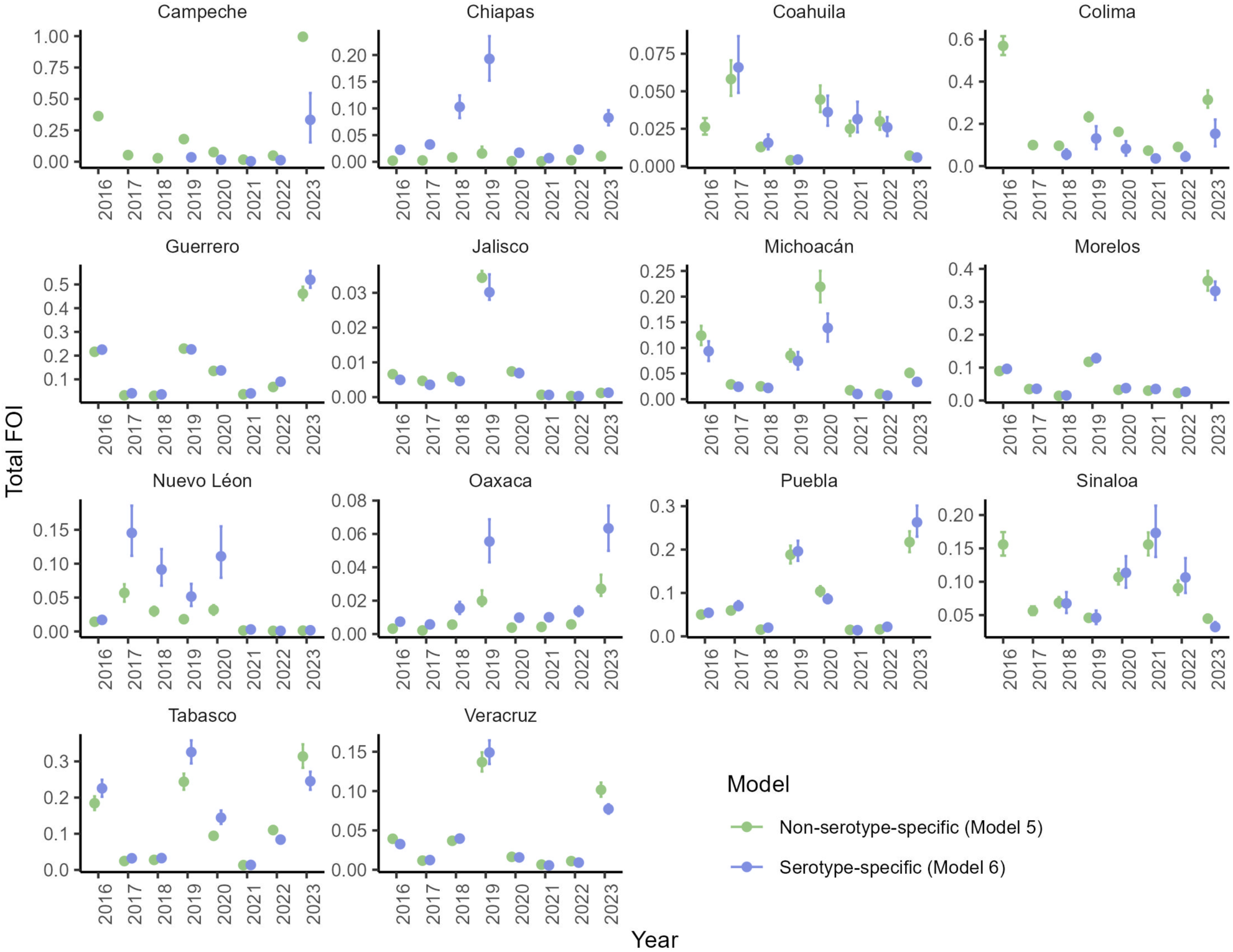
Comparison of model output from Models 5 and 6. Median overall FOI estimates and 95% credible intervals generated by the non-serotype-specific model (Model 5, green) and the serotype-specific model (Model 6, blue) for states modelled for more than five years in Model 6 (and also for Campeche, which was modelled for five years).

## Discussion

This study shows that both overall and serotype-specific dengue FOIs vary substantially in time across Mexico. The serotype-specific model developed in this paper gives new insights on the transmission intensity of DENV serotypes across Mexico, challenging the assumption of equal transmissibility of the four serotypes. We found substantial regional heterogeneity in dengue incidence and transmission intensity in Mexico between 2016-2023, and over the 8-year period, DENV-2 was the most frequently detected serotype, followed by DENV-1. DENV-3 frequency was low at the start of the reporting period but substantially increased between 2021 and 2023 in many states in the south-east of the country, and few DENV-4 cases were reported across the whole study period.

In 2023, one of the worst years for dengue in Mexico in the last decade^(25)^, different DENV serotypes caused simultaneous outbreaks in different parts of the country; in the south-east of Mexico, notable outbreaks in Campeche, Tabasco and Quintana Roo were driven by DENV-3, for which we estimated median FOIs of 0.3311 (95% CrI 0.1526-0.5420), 0.2059 (95% CrI 0.1859-0.2278), and 0.1047 (95% CrI 0.0319-0.5814), respectively, and in the centre and west of the country, a resurgence of DENV-1 transmission led to the substantial rise in cases in Puebla (median DENV-1 FOI: 0.1902 [95% CrI 0.1560-0.2264]), and Guerrero (median DENV-1 FOI: 0.2592 [95% CrI 0.2313-0.2900]).

The non-serotype-specific FOI estimates from the constant-in-time non-serotype-specific model (Model 1) range from 0.0366 (95% CrI 0.0345-0.0389) to 0.0727 (95% CrI 0.0717-0.0736), which spans a similar though slightly narrower range than the estimates generated in Rodriguez-Barraquer et al.^(18)^. Much like in O’Driscoll et al.^(16)^, we found that the modelled time-varying non-serotype specific FOIs (i.e. those generated using Model 5) were more variable than the constant-in-time non-serotype specific FOIs generated using Model 1. Our constant-in-time FOI estimates tended to be higher in states in the south-east of Mexico than in other states. This finding is also borne out in the estimates generated from the time-varying (non-serotype-specific) Model 5, with high dengue FOI estimates in 2023 in states such as Yucatán and Campeche. The estimated FOI in Yucatán from the constant-in-time non-serotype-specific model (Model 1) (0.0727 [95% CrI 0.0717-0.0736]) is higher than (but within the credible interval of) the estimate generated in Vicco et al.^(26)^ from the analysis of seroprevalence data from 2014 (0.045 [95% CrI 0.020-0.081]). The estimated FOI in Morelos from Model 1 (0.0473 [95% CrI 0.0465-0.0480]) is also similar to the estimate generated in Vicco et al.^(26)^ from seroprevalence data from 2011 (0.044 [95% CrI 0.018-0.087]).

Both the serotype-specific and non-serotype-specific models ignore cross-protection, assume endemic transmission of dengue prior to the first modelled year (2016), via a constant historical FOI estimate, and may also be limited by modelling reporting rates as constant in time and by serotype, as increased public awareness during outbreak years may increase reporting rates and previous serotype-specific modelling work suggests that reporting rates may differ between serotypes^(27)^. It is interesting that the performance of the time-varying non-serotype-specific models, as assessed by the DIC, improved when we assumed that the probability of reporting a dengue infection among children under 15 years of age was higher than among the rest of the population, which has been noted in Ecuador^(28)^. However, the extent to which children reported more or less than adults differed by state, and more work can be done in the future to better understand age trends in dengue case reporting rates.

The serotype-specific version of the model, whilst being an improvement over the non-serotype-specific versions, is limited by data availability. Given only 10.3% of cases had a positive RT-PCR result, we reconstructed serotype-specific case data by scaling the total number of hospitalised and non-hospitalised cases by the respective proportions of each serotype observed from RT-PCR testing. As such, these reconstructed data have limitations, as we could not correct for potential differences in serotype-specific severity (as measured by the propensity of hospitalisation) and the propensity to be RT-PCR-tested (which was greater in hospitals). Additionally, we assumed that a serotype was not co-circulating if it had fewer than 10 cases detected during every year of data, which could be affected by different propensities to cause severe disease, hospitalisation, and RT-PCR-testing. This assumption also meant that if a serotype was not circulating between 2016-2023, we assumed that it was also not circulating in the years before 2016, so any previous contribution of that serotype’s historical circulation to population immunity profiles was not accounted for in the model, possibly biasing the initial susceptibility of the population and hence the FOI estimates generated.

More generally, the surveillance system in Mexico has limitations, as is the case with all passive epidemiological surveillance systems^(29)^. In particular, there are differences between hospitalised and ambulatory cases, with underreporting higher in the latter group^(30)^. The database used in this paper is curated and validated annually by the health authorities, however, since transmission intensity changes annually, there may be annual variations in the magnitude of under-reporting. Only 30% of probable outpatient dengue cases are laboratory confirmed during epidemic peaks in Mexico^(31)^, and since the criterion is subjective (there is no definition of an “epidemic peak”), there is no way to quantify the magnitude of this under-reporting. Finally, medical clinics associated with pharmacies (CAP) have expanded in Mexico since 2010; according to the 2023 National Health and Nutrition Survey^(32)^, the number of consultations in this type of primary care clinic could reach 20% of all medical consultations nationwide. In theory, all providers of medical consultations are required to report probable cases of dengue. However, in practice, these CAPs do not report to the surveillance system. These limitations may affect the calculations presented here; however, they do not diminish the magnitude of dengue transmission in Mexico as shown in this study.

Increased RT-PCR-testing in Mexico and across the world would help generate more accurate transmission intensity estimates for the different serotypes. Our estimates suggest that DENV-4 had the lowest FOI of all four serotypes in the eight states in which we modelled DENV-4 transmission (Figure 6 and Figures S10, S13, S20, S22, S26-28, Supplementary Information). Serotype-specific seroprevalence data would be particularly useful to validate the estimated serotype-specific trends in FOI, as these are not consistent with the serotype-specific FOI estimates obtained in Iquitos (Peru)^(33)^ and in French Polynesia^(27)^.

Serotype-specific seroprevalence studies would be extremely useful to disentangle the potential mechanisms behind the heterogeneities in serotype circulation. The development of mathematical models to estimate the serotype-specific transmission intensity of dengue when serotypes are co-circulating from routine case data is a novel contribution of this paper which we hope will stimulate new research and evidence generation from multiple settings. Better characterising serotype-specific variations in transmissibility will allow the generation of refined global DENV transmission intensity and burden maps (such as https://arbomap.org), which currently assume equal transmission intensity of the four serotypes. Most importantly, given all currently available (and most likely also upcoming) dengue vaccines have complex and serotype-specific efficacy profiles, it is crucial to assess the extent to which the different serotypes may differ in their transmission potentials and in their propensities to generate severe disease, as these estimates will inform vaccine impact modelling studies, global guidelines and national vaccination programmes going forward.

## Data Availability

Data and code used to generate results is publicly available and can be accessed through GitHub, via the url https://github.com/mrc-ide/Overall-and-Serotype-Specific-Dengue-Transmission-Trends-in-Mexico-2016-2023.

## References

1. Pan American Health Organization. Dengue: analysis by country 2025 [Available from: https://www.paho.org/en/arbo-portal/dengue-data-and-analysis/dengue-analysis-country.

2. Cattarino L, Rodriguez-Barraquer I, Imai N, Cummings DAT, Ferguson NM. Mapping global variation in dengue transmission intensity. Science Translational Medicine. 2020;12(528):eaax4144.

3. Colón-González FJ, Sewe MO, Tompkins AM, Sjödin H, Casallas A, Rocklöv J, et al. Projecting the risk of mosquito-borne diseases in a warmer and more populated world: a multi-model, multi-scenario intercomparison modelling study. Lancet Planet Health. 2021;5(7):e404–e14.

4. United Nations Department of Economic and Social Acairs PD. World Urbanization Prospects 2018, Country Profiles 2018 [Available from: https://population.un.org/wup/Country-Profiles/.

5. National Institute of Statistics and Geography (INEGI). Census of Population and Housing 2020 [Available from: https://en.www.inegi.org.mx/programas/ccpv/2020/.

6. Dantés HG, Farfán-Ale JA, Sarti E. Epidemiological Trends of Dengue Disease in Mexico (2000–2011): A Systematic Literature Search and Analysis. PLOS Neglected Tropical Diseases. 2014;8(11):e3158.

7. Lozano-Fuentes S, Hayden MH, Welsh-Rodriguez C, Ochoa-Martinez C, Tapia-Santos B, Kobylinski KC, et al. The dengue virus mosquito vector Aedes aegypti at high elevation in Mexico. Am J Trop Med Hyg. 2012;87(5):902–9.

8. Colón-González FJ, Fezzi C, Lake IR, Hunter PR. The Ecects of Weather and Climate Change on Dengue. PLOS Neglected Tropical Diseases. 2013;7(11):e2503.

9. Johansson MA, Cummings DAT, Glass GE. Multiyear Climate Variability and Dengue—El Niño Southern Oscillation, Weather, and Dengue Incidence in Puerto Rico, Mexico, and Thailand: A Longitudinal Data Analysis. PLOS Medicine. 2009;6(11):e1000168.

10. Ferguson N, Anderson R, Gupta S. The ecect of antibody-dependent enhancement on the transmission dynamics and persistence of multiple-strain pathogens. Proc Natl Acad Sci U S A. 1999;96(2):790–4.

11. Reich NG, Shrestha S, King AA, Rohani P, Lessler J, Kalayanarooj S, et al. Interactions between serotypes of dengue highlight epidemiological impact of cross-immunity. J R Soc Interface. 2013;10(86):20130414.

12. Katzelnick LC, Gresh L, Halloran ME, Mercado JC, Kuan G, Gordon A, et al. Antibody-dependent enhancement of severe dengue disease in humans. Science. 2017;358(6365):929–32.

13. Sarti E, L’Azou M, Mercado M, Kuri P, Siqueira JB, Solis E, et al. A comparative study on active and passive epidemiological surveillance for dengue in five countries of Latin America. International Journal of Infectious Diseases. 2016;44:44–9.

14. Ferguson NM, Donnelly CA, Anderson RM. Transmission dynamics and epidemiology of dengue: insights from age-stratified sero-prevalence surveys. Philos Trans R Soc Lond B Biol Sci. 1999;354(1384):757–68.

15. Imai N, Dorigatti I, Cauchemez S, Ferguson NM. Estimating Dengue Transmission Intensity from Case-Notification Data from Multiple Countries. PLOS Neglected Tropical Diseases. 2016;10(7):e0004833.

16. O’Driscoll M, Imai N, Ferguson NM, Hadinegoro SR, Satari HI, Tam CC, et al. Spatiotemporal variability in dengue transmission intensity in Jakarta, Indonesia. PLOS Neglected Tropical Diseases. 2020;14(3):e0008102.

17. Imai N, Dorigatti I, Cauchemez S, Ferguson NM. Estimating Dengue Transmission Intensity from Sero-Prevalence Surveys in Multiple Countries. PLOS Neglected Tropical Diseases. 2015;9(4):e0003719.

18. Rodriguez-Barraquer I, Salje H, Cummings DA. Opportunities for improved surveillance and control of dengue from age-specific case data. eLife. 2019;8:e45474.

19. Dirección General de Epidemiología SdS. Datos Abiertos Dirección General de Epidemiología 2024 [Available from: https://www.gob.mx/salud/documentos/datos-abiertos-152127

20. National Institute of Statistics and Geography (INEGI). Intercensal Survey 2015 [Available from: https://en.www.inegi.org.mx/programas/intercensal/2015/.

21. Spiegelhalter DJ, Best NG, Carlin BP, Van Der Linde A. Bayesian Measures of Model Complexity and Fit. Journal of the Royal Statistical Society Series B: Statistical Methodology. 2002;64(4):583–639.

22. St. John AL, Rathore APS. Adaptive immune responses to primary and secondary dengue virus infections. Nature Reviews Immunology. 2019;19(4):218–30.

23. Stan Develpment Team. Stan Modeling Language Users Guide and Reference Manual. 2.35 ed2024.

24. R Core Team. R: A Language and Environment for Statistical Computing. 4.4.0 ed2024.

25. Pan American Health Organization. Reported Cases of Dengue Fever in The Americas [Available from: https://www3.paho.org/data/index.php/en/mnu-topics/indicadores-dengue-en/dengue-nacional-en/252-dengue-pais-ano-en.html.

26. Vicco A, McCormack C, Pedrique B, Ribeiro I, Malavige GN, Dorigatti I. A scoping literature review of global dengue age-stratified seroprevalence data: estimating dengue force of infection in endemic countries. eBioMedicine. 2024;104:105134.

27. Nemoto T, Aubry M, Teissier Y, Paul R, Cao-Lormeau V-M, Salje H, et al. Reconstructing long-term dengue virus immunity in French Polynesia. PLOS Neglected Tropical Diseases. 2022;16(10):e0010367.

28. Vitale M, Lupone CD, Kenneson-Adams A, Ochoa RJ, Ordoñez T, Beltran-Ayala E, et al. A comparison of passive surveillance and active cluster-based surveillance for dengue fever in southern coastal Ecuador. BMC Public Health. 2020;20(1):1065.

29. Murray J, Cohen AL. Infectious Disease Surveillance. In: Quah SR, editor. International Encyclopedia of Public Health (Second Edition). Oxford: Academic Press; 2017. p. 222–9.

30. Undurraga EA, Betancourt-Cravioto M, Ramos-Castañeda J, Martínez-Vega R, Méndez-Galván J, Gubler DJ, et al. Economic and Disease Burden of Dengue in Mexico. PLOS Neglected Tropical Diseases. 2015;9(3):e0003547.

31. Dirección General de Epidemiología. Manual de Procedimientos Estandarizados para la Vigilancia Epidemiológica de las Enfermedades Transmitidas por Vector (ETV). In: Secretaría de Salud SadPnyPndlS, editor. 2021.

32. Centro de Investigación en Evaluación y Encuestas (CIEE). Encuesta Nacional de Salud y Nutrición Continua 2023 [Available from: https://ensanut.insp.mx/encuestas/ensanutcontinua2023/descargas.php.

33. Reiner RC, Stoddard ST, Forshey BM, King AA, Ellis AM, Lloyd AL, et al. Time-varying, serotype-specific force of infection of dengue virus. Proceedings of the National Academy of Sciences. 2014;111(26):E2694–E702.

